# Use of Medroxyprogesterone Acetate and Risk of Meningiomas: A Comparative Safety Study

**DOI:** 10.1101/2025.01.25.25321119

**Authors:** Connor Frey, Mohit Sodhi, Mostafa Fatehi, Abbas Kezouh, Mahyar Etminan

## Abstract

Medroxyprogesterone acetate (MPA), a popular contraceptive agent has recently been associated with meningiomas. However, this risk compared to other available contraceptive agents has not been elucidated.

We sought to quantify the risk of meningiomas in women on MPA compared to those using oral contraceptives containing ethinylestradiol-levonorgestrel (EE-LNG) by undertaking a nested case-controls study using the PharMetrics Plus database (IQVIA, USA). After creating a cohort of new users of MPA and EE-LNG from 2006 to 2020 new cases of meningioma were identified. Cases were and matched to controls by age and calendar time. Conditional logistic regression was used to adjust for previous exposure to radiotherapy, previous use of hormonal contraceptives, and obesity The adjusted incident rate ratio (IRR) for meningioma for greater than one year of use was 3.55 (95%ICI: 1.85-6.55). Women using MPA have an increased risk of developing meningioma compared to users of EE-LNG.

---

Medroxyprogesterone acetate (MPA, Depo-Provera®) is a popular hormonal contraceptive administered intramuscularly used by women around the world, with approximately one million dispensed prescriptions annually in the United States^1^. MPA is a popular alternative for women who are less adherence to oral contraceptives as it is administered every 90 days.

A recent nested case-control study has shown that MPA increases the risk of meningioma by nearly six-fold (OR= 5.62, 95%CI:2.19-14.42)^2^. However, this risk with MPA was based only on 8 exposed cases and did not adequately control for disease latency. Moreover, given the wide use of hormonal contraceptives by women worldwide, the risk of meningioma with MPA must be compared to an active comparator. Meningiomas are considered latent outcomes, and failure to control for disease latency can lead to reverse causation where a meningioma could have occurred prior to the use of the drug.

Meningiomas make up 40% of central nervous system tumors and can range from low grade benign tumors to higher grade malignant tumours. In light of the popularity of MPA among women worldwide, the risk of meningioma with this popular medication needs to be better quantified and compared to other alternative contraceptive medications allowing women to make more informed decisions when deciding to take this class of medications.

We used the PharMetrics^®^ Plus for Academics, a health plan claims database comprised of fully adjudicated medical and pharmacy claims for more than 114 million unique enrollees since 2006 with fully adjudicated pharmacy and medical claims^3^. The medical claims data is comprised of all physician visits which captures medical diagnoses through international classification of diseases, ninth and tenth editions (ICD-9 and 10). All prescription drug data includes information on drug name, dose quantity dispensed, and day supply. Ethics approval was obtained from the University of British Columbia Clinical Ethics Board.

We created a cohort of new users of MPA and a control cohort of oral ethinylestradiol- levonorgestrel (EE-LNG) users. We allowed a one-year look back period to ensure study drugs are incident users and also used this period for covariate adjustment.

The cohort was followed to the first incidence of meningioma (both intracranial and spinal) ascertained by ICD-9 or ICD-10. The date of each case was deemed the index date. For each case, a risk-set of all controls who had the same follow up time to that of the case were identified. From each risk set we selected 4 controls using a density-based sampling approach where a control could become a future case. Cases and controls were matched by age (±1 year) and calendar time. This manner of control selection has shown to generate odds ratios that are close approximation to the incident rare ratio (IRR)^4^.

MPA or EE-LNG exposure was defined as use of at least two prescriptions of a study drug two years from the index date where the first year prior to the index date (day 366-730) was disregarded and designated a grace period to control for disease latency (greater than one year exposure). As a sensitivity analysis, we further incorporated longer lag periods where the first two years, three years and four years prior to the index date were lagged allowing for greater than two, three and four years of exposure. Conditional logistic regression was used to estimate IRRs, adjusting for previous use of radiation therapy, use of other contraceptives, and obesity.

The cohort consisted of 72,181 MPA users and 247,180 EE-LNG users. Among the cohort, we identified 212 cases and 848 controls. The average age for cases and controls was 40.8 (± 9.8) The distribution of obesity, previous radiation therapy and use of previous contraceptives was comparable between the cases and controls (Table 1).

**Table 1:** Demographics of cases and controls.

|  | **Cases** | **Controls** |
| --- | --- | --- |
| **Number** | 212 | 848 |
| **Age (in years, mean ± SD)** | 40.8 ± 9.8 | 40.8 ± 9.7 |
| **Obesity (%)** | 3.3 | 6.3 |
| **Previous radiation therapy (%)** | 2.8 | 3.0 |
| **Previous use of contraceptives** | 19.3 | 22.3 |

Overall, the crude and adjusted IRRs were similar. The adjusted odds ratio for greater than one year of use was 3.55 (95%ICI: 1.85-6.55) (Table 2). The adjusted IRRs for greater than 2,3 and 4 years of use were 2.95 (95%CI: 1.68-5.18), 3.17 (95%CI:1.71-5.88) and 3.50 (95% CI:1.70-7.19) respectively (Table 2).

**Table 2.**
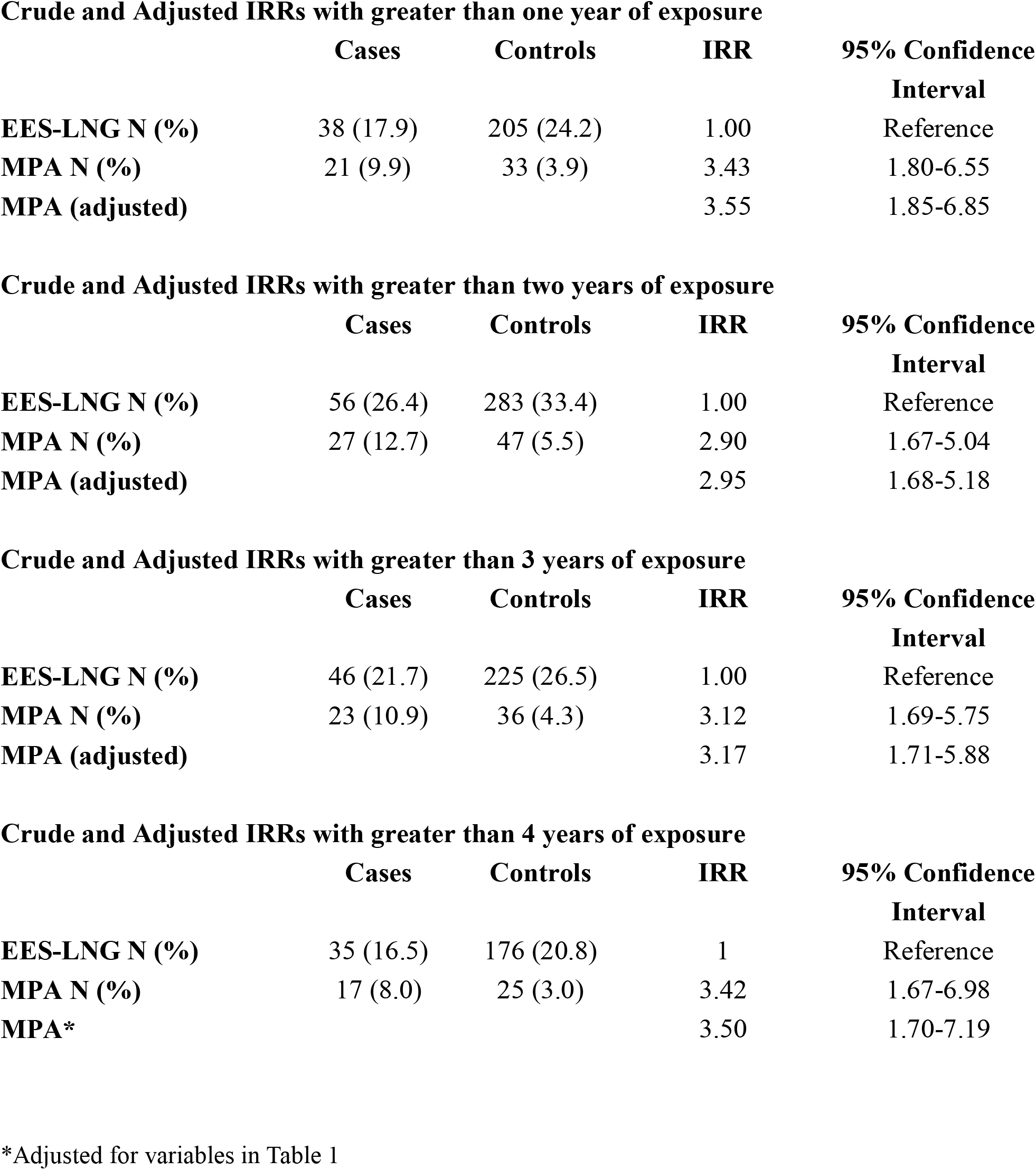
Crude and adjusted incident rate ratios (IRRs) for medroxyprogesterone (MPA) and ethinylestradiol-levonorgestrel (EES-LNG) with varying washout periods.

The results of our study suggest an increase in the risk of meningiomas when compared to the more commonly prescribed oral contraceptives containing EE-LNG. The strength of our study is the tight control for disease latency which is critical for this question. Failure to control for the long latency of meningioma can lead to reverse causation where the onset of the disease could have occurred prior to the use of the drug.

The 3.5 increase in risk among MPA users needs to be put into perspective. The annual incidence of meningiomas among women in the United States is estimated to be 12/100,000. Assuming a constant risk increase, the three-year cumulative risk would be 36/100,000. (0.0365) A 3.5 higher risk with MPAs (compared to EE-LNG) would translate to a new risk of 126/100,000 (0.126%) and an absolute risk increase of 0.09% (0.126%-0.0365%). The reciprocal of the difference (1/0.09%) is the number needed to harm^5^ which would be 1,111. This means that for every 1,111 women on MPA (for three years), one would experience a meningioma.

Two epidemiologic studies to date have examined the risk of meningiomas with MPA. The nested case-control study by Roland found a 5.6 increase in risk of meningiomas with MPA^2^. However, the study did not control for disease latency, did not have an active comparator, and only had 8 exposed cases. Another case-control study by Griffin et al. also found an increase in risk of 2.86 (1.79–4.57) for 2-3 years of use. Similarly, this study also did not control for disease latency and did not have an active control group^6^.

The proposed mechanism by which MPA may contribute to meningioma development involves its stimulation of tumour cell proliferation via hormone receptor binding, particularly to progesterone receptors, which are commonly overexpressed in meningiomas^7^. Activation of these receptors initiates signalling pathways that enhance cellular proliferation and survival, creating an environment conducive to tumour growth.

Recently, the European Medicines Agency has recommended addition of meningioma as a warning in the drug’s label^8^. However, to date, a label change has not taken place by the Food and Drug Administration. Our study adds to the existing body of evidence that MPA can increase the risk of meningioma. We believe the results of this study further warrants the need to alert women about this serious, albeit rare adverse event.

The strength of our study is the large sample size and optimal control for disease latency, going back up to 4 years from the date of diagnosis. Moreover, using a popular comparator group of EE-LNG users provided a more homogenous comparison (as opposed to nonusers of any contraceptives), allowing women to compare the risks associated with MPA to another commonly used contraceptive.

The results of this study suggest and increase in the risk of meningiomas with MPA use compared to EE-LNG. Women who are concerned about this risk can opt to taking oral contraceptives containing EE-LNG.

## Data Availability

Some of the data may be available upon request

## Data Availability Statement/Data Sharing Plan

The data are not publicly available because of privacy restrictions set by the data owners, IQVIA (USA).

## Conflict of Interest Disclosures

None declared.

## Funding/Support

This study was funded by internal research funds from the Department of Ophthalmology and Visual Sciences, University of British Columbia.

## Notes

### Competing Interest Statement

The authors have declared no competing interest.

### Funding Statement

This study did not receive any funding

